# Dynamics and significance of the antibody response to SARS-CoV-2 infection

**DOI:** 10.1101/2020.07.18.20155374

**Authors:** Anita S. Iyer, Forrest K. Jones, Ariana Nodoushani, Meagan Kelly, Margaret Becker, Damien Slater, Rachel Mills, Erica Teng, Mohammad Kamruzzaman, Wilfredo F. Garcia-Beltran, Michael Astudillo, Diane Yang, Tyler E. Miller, Elizabeth Oliver, Stephanie Fischinger, Caroline Atyeo, A. John Iafrate, Stephen B. Calderwood, Stephen A. Lauer, Jingyou Yu, Zhenfeng Li, Jared Feldman, Blake M. Hauser, Timothy M. Caradonna, John A. Branda, Sarah E. Turbett, Regina C. LaRocque, Guillaume Mellon, Dan H. Barouch, Aaron G. Schmidt, Andrew S. Azman, Galit Alter, Edward T Ryan, Jason B. Harris, Richelle C. Charles

**Affiliations:** Division of Infectious Diseases, Massachusetts General Hospital, Boston, MA, USA; Department of Medicine, Harvard Medical School, Boston, MA, USA; Department of Epidemiology, Johns Hopkins Bloomberg School of Public Health, Baltimore, MD, USA; Department of Pathology, Massachusetts General Hospital, Boston, MA, USA; Ragon Institute of MGH, MIT, and Harvard, Cambridge, MA, USA; Department of Microbiology, Harvard Medical School, Boston, MA, USA; Center for Virology and Vaccine Research, Beth Israel Deaconess Medical Center, Harvard Medical School, Boston, MA, USA; Department of Immunology and Infectious Diseases, Harvard T.H. Chan School of Public Health, Boston, MA, USA; Department of Pediatrics, Harvard Medical School, Boston, MA, USA

## Abstract

**BACKGROUND:** Characterizing the humoral immune response to SARS-CoV-2 and developing accurate serologic assays are needed for diagnostic purposes and estimating population-level seroprevalence.

**METHODS:** We measured the kinetics of early antibody responses to the receptor-binding domain (RBD) of the spike (S) protein of SARS-CoV-2 in a cohort of 259 symptomatic North American patients infected with SARS-CoV-2 (up to 75 days after symptom onset) compared to antibody levels in 1548 individuals whose blood samples were obtained prior to the pandemic.

**RESULTS:** Between 14-28 days from onset of symptoms, IgG, IgA, or IgM antibody responses to RBD were all accurate in identifying recently infected individuals, with 100% specificity and a sensitivity of 97%, 91%, and 81% respectively. Although the estimated median time to becoming seropositive was similar across isotypes, IgA and IgM antibodies against RBD were short-lived with most individuals estimated to become seronegative again by 51 and 47 days after symptom onset, respectively. IgG antibodies against RBD lasted longer and persisted through 75 days post-symptoms. IgG antibodies to SARS-CoV-2 RBD were highly correlated with neutralizing antibodies targeting the S protein. No cross-reactivity of the SARS-CoV-2 RBD-targeted antibodies was observed with several known circulating coronaviruses, HKU1, OC 229 E, OC43, and NL63.

**CONCLUSIONS:** Among symptomatic SARS-CoV-2 cases, RBD-targeted antibodies can be indicative of previous and recent infection. IgG antibodies are correlated with neutralizing antibodies and are possibly a correlate of protective immunity.

## INTRODUCTION

Severe acute respiratory syndrome coronavirus 2 (SARS-CoV-2), the causative agent of coronavirus disease 2019 (COVID-19), has spread rapidly around the world since first identified in Wuhan, China, in December 2019^1^. On March 11^th^, 2020 the World Health Organization (WHO) declared COVID-19 a pandemic. As of July 13th, 2020, the disease has caused 12,750,275 confirmed cases and 566,355 deaths globally^2^.

Currently, our understanding of antibody responses following infection with SARS-CoV- 2 is limited^3,4^, including the magnitude and duration of responses, cross-reactivity with other coronaviruses and viral respiratory pathogens, and correlates of protective immunity following infection. A detailed characterization of antibody responses is needed to determine whether antibody-based tests can augment viral detection-based assays in the diagnosis of active or recent infections and to inform the design and interpretation of seroepidemiologic studies.

In this study, we characterize the kinetics and antibody isotype profile to the receptor binding domain (RBD) of the spike (S) protein of SARS-CoV-2 in a longitudinal cohort of North American patients infected with SARS-CoV-2 and in pre-pandemic controls. We evaluated the sensitivity and specificity of anti-RBD responses in detecting recent infection and estimated the time it takes for cases to become seropositive (seroconversion) or return to seronegative (seroreversion). We also examined how well these responses correlated with neutralizing antibody activity directed at the S protein. Additionally, we evaluated the cross-reactivity of these responses with other coronavirus RBDs and characterize assay performance using dried blood spots.

## MATERIALS/ METHODS

### Plasma/serum/dried blood spot (DBS) samples

Clinical samples were obtained from individuals with PCR confirmed SARS-COV-2 infection presenting to the Massachusetts General Hospital (MGH) in Boston, MA with fever and/or viral respiratory symptoms from March to April 2020 and who met criteria for RT-PCR testing. These criteria changed over time, but included patients with severe symptoms requiring hospital admission, who had other risk factors for disease progression (e.g. were age 60 or older, had diabetes, or were immunocompromised), or who worked or lived in a setting where infection control requirements dictated a need for testing. Additional serum/plasma samples collected September 2015 to December 2019, prior to the SARS-COV-2 outbreak, were used as controls. This included healthy adults seen at the MGH Immunization and Travel Clinic prior to travel, patients undergoing routine serology, and patients presenting with other known febrile illnesses. Plasma samples, except for the routine serology samples, were heat-inactivated at 56°C for one hour prior to analysis. DBS sample preparation is provided in the Supplementary Material. Patient demographic information, lab results, and clinical outcomes were extracted from the electronic medical record. All research was approved by the Institutional Review Board for Human Subjects Research at MGH.

### Enzyme-linked immunosorbent assay (ELISA)

The ELISA assays measured IgG, IgA, and IgM responses to the receptor binding domain of the spike protein (RBD) from SARS-CoV-2 [GenBank: MN975262], Middle East Respiratory Syndrome (MERS) virus [GenBank: AFY13307.1], SARS-CoV-1 [GenBank: AAP13441.1], and common cold coronaviruses HKU1 [GenBank: AAT98580.1], OC229E [GenBank: AAK32191], OC43 [GenBank:AAT84362], and NL63 [GenBank: AKT07952]. Anti**-** RBD-specific antibody concentrations (ug/mL) were quantified using isotype-specific anti-RBD monoclonal antibodies^5^. A full protocol is provided in the Supplementary Material and is available on protocols.io (https://www.protocols.io/view/sars-cov-2-rbd-elisa-bikbkcsn).

### Pseudovirus neutralization assay

To determine the SARS-CoV-2 neutralization activity of our plasma samples, we used a lentivirus pseudoneutralization model as previously described^6^, which is a strong correlate of protective immunity in challenged rhesus macaques^7^. We expressed results from this assay as the antibody titer required to neutralize 50% of the SARS-CoV-2 pseudovirus (NT50).

### Statistical analysis

#### Single isotype thresholds

We first explored how cutoffs of individual isotypes (IgM, IgG and IgA) performed in identifying previously infected individuals. We compared measurements from pre-pandemic controls, with those taken at any time, ≤7 days, 8-14 days,>15-28 days, and >28 days after the onset of symptoms. Using a previously described cross-validation procedure^8^, we allocated both cases and controls among 10-folds to calculate a pooled cross-validated AUC (cvAUC). Using the isotype cut-offs defined by the maximum concentration (ug/mL) found among pre-pandemic controls (IgG: 0.57, IgM: 2.63, IgA: 2.02), we estimated sensitivity and bootstrap 95% confidence intervals.

#### Random forest classification models

We explored how combining multiple isotype-specific responses with a random forest classification models performed identifying previously infected individuals. We repeated the same procedures for cross-validation and assessed variable importance within these different models using a permutation test–based metric, mean decrease in accuracy.

#### Analysis of time to seroconversion and seroreversion

Using the cut-offs defined earlier, we modeled the time required to become seropositive (seroconversion) and return to becoming seronegative (seroreversion). Using individual level interval-censored data, we fit non-parametric (i.e Turnbull’s Estimator) and parametric accelerated failure time models using the icenReg R package^9^. All time-to-event data were assumed to be Weibull distributed with bootstrapped 95% confidence intervals estimated.

All analyses were completed using R (Version 3.6.1) within Rstudio (Version 1.2.5019).

## RESULTS

### Study cohorts

We measured RBD responses in two cohorts: 1) symptomatic patients who tested positive for SARS-CoV-2 by PCR (n = 259) and 2) healthy (n = 1,515) and febrile controls (n = 33) collected prior to the SARS-CoV-2 outbreak. The majority of SARS-CoV-2 positive cases were severe (95% hospitalized, 31% requiring ICU level care, 16% died), male (62%), and older (median age: 58) (Table 1, Figure S1). Most pre-pandemic controls were younger (median age: 37) and female (62%). Serum was collected at multiple time points for most patients (56%; n=146), with 23% (n=59) having ≥4 samples. Forty-nine percent of this cohort had a sample collected between 0-7 days after onset of symptoms (n=126), 59% had a sample between 8-14 days (n=154), 42% had a sample between 15-28 days (n=110), 22% had a sample between 29-45 days (n=56), 8% had a sample between 46-60 days (n=20) and 3% had a sample > 60 days (n=7). The last sample was collected 75 days post-symptom onset.

**Table 1.** Individual characteristics of PCR-positive SARS-CoV-2 cases and pre-pandemic controls.

| Characteristic | Pre-pandemic Controls*<br>(N=1,548) | PCR-positive Cases<br>(N=259) |
| --- | --- | --- |
| Age |  |  |
| Median [IQR] | 37 [30-54] | 58 [44-72] |
| <65 years (%) | 1,386 (90) | 160 (62) |
| 65+ years (%) | 162 (10) | 99 (38) |
| Female (%) | 1,024 (66) | 98 (38) |
| Immunocompromised (%) | NA | 17 (7) |
| Severity |  |  |
| Not Hospitalized (%) | NA | 14 (5) |
| Hospitalized, no ICU (%) | NA | 122 (47) |
| Hospitalized, required ICU (%) | NA | 81 (31) |
| Died due to COVID-19 (%) | NA | 41 (16) |
| Missing Severity (%) | NA | 1 (0) |

\*Pre-pandemic controls included healthy adults (n=278), patients undergoing routine serology testing (n=1241), and patients presenting with other known febrile illnesses (n = 33), including 13 with bacteremia (e.g. *S. aureus*, *S. pneumoniae*, *E. coli*, or *K. pneumoniae* confirmed by standard microbiologic techniques), 4 with babesiosis (confirmed by microscopy and/or PCR), 1 with presumed scrub typhus, and 15 with viral respiratory infections (e.g. influenza [7], parainfluenza [4], respiratory syncytial virus [3], and metapneumovirus [1] confirmed by PCR or direct fluorescent antibody test

### Kinetics of anti-SARS-CoV-2 RBD antibody responses

Most cases eventually had evidence of elevated antibodies to SARS-CoV-2 compared to pre-pandemic controls (Figure 1). From days 5 to 14, there was a sharp rise in RBD-specific antibodies of all isotypes, and IgG titers continued to rise until day 25 after the onset of symptoms (Figure S2A). The population average IgA and IgM responses peaked a few days earlier than IgG and then declined towards levels measured in pre-pandemic samples (Figure S2 and S3). IgG antibody responses also began to wane, but at a slower rate. Among 58 cases with ≥4 measurements, the individual peak measurement IgM often occurred before that of IgG (Before: 48%, Same: 48%) and simultaneously with that of IgA (Before: 22%, Same: 62%). Among hospitalized patients, the population average trajectory differed little between severity levels; the average IgM levels among hospitalized cases admitted to the ICU dropped more quickly than that of patients not requiring the ICU or who died (Figure S2B).

**Figure 1.**
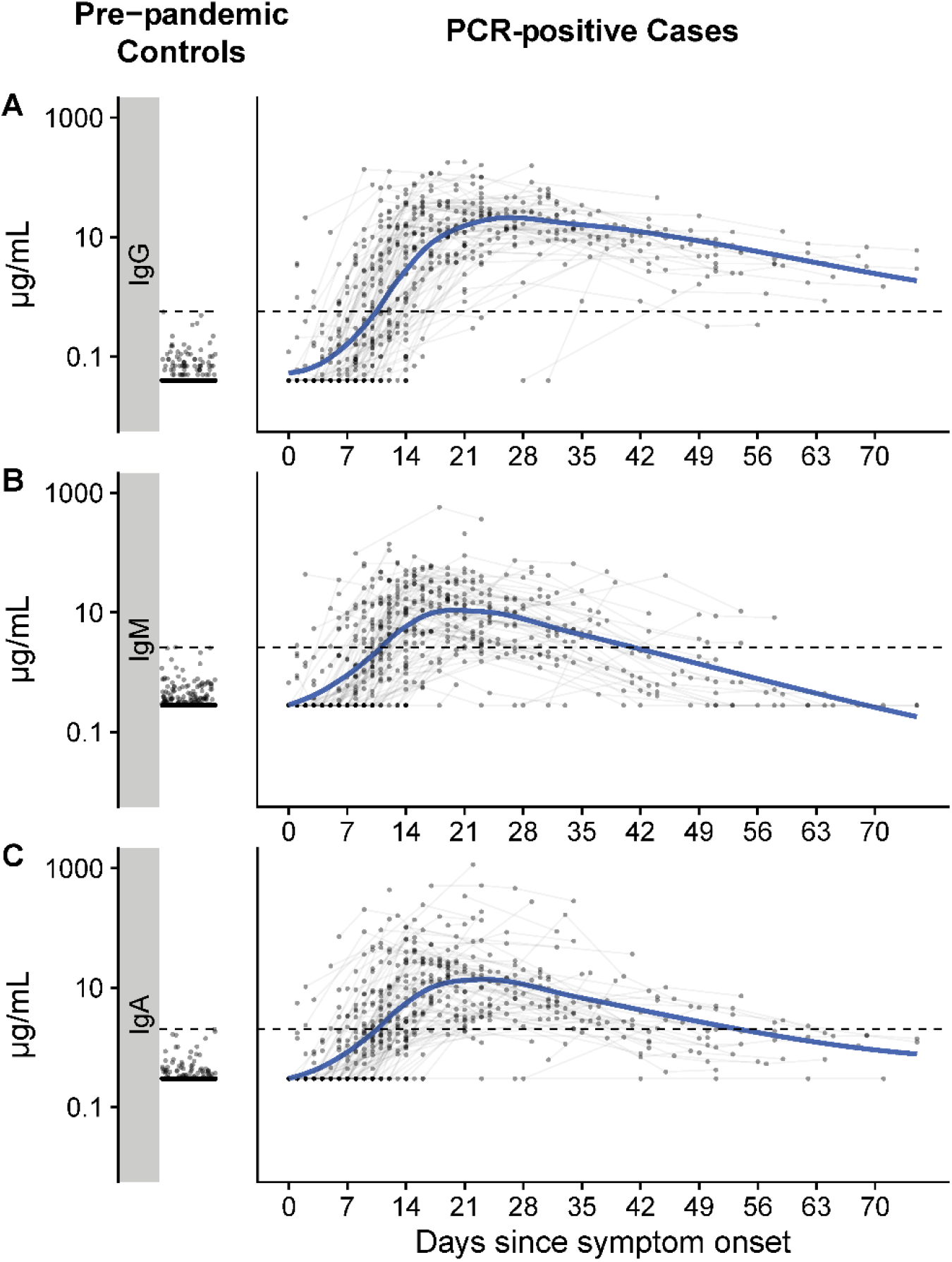
Measurements of IgG, IgM, and IgA against SARS-CoV-2 spike protein receptor binding domain among pre-pandemic controls and PCR positive cases. Each dot represents a unique measurement of an isotype (Row A: IgG, Row B: IgM, Row C: IgA) in pre-pandemic controls (left panels) and PCR positive cases (right panels). The blue line is a loess smooth non- parametric function. Black dashed lines indicate the maximum concentration (µg/mL) found among pre-pandemic controls (IgG: 0.57, IgM: 2.63, IgA: 2.02). Horizontal jitter was introduced into the pre-pandemic controls. The limit of detection (µg/mL) was 0.04 for IgG, 0.28 for IgM, and 0.3 for IgA.

### Accuracy of RBD antibodies for identifying recent SARS-CoV-2 infection

Each antibody isotype was predictive of infection, and the cross-validated AUC (cvAUC) of antibody testing for each antibody isotype increased to above 97% during the period of 15-28 days after symptom onset (Table 2). The cvAUC remained high for IgG after 28 days but began to fall for IgM and IgA. Using the pre-defined cutoffs, the sensitivity of IgG antibodies rose from 7% (<=7days) to 97% after 14 days of symptoms. The sensitivity of IgA and IgM rose to 91% and 81% 2-4 weeks post-symptom onset but dropped after 4 weeks to 57% and 40%, respectively.

**Table 2.**
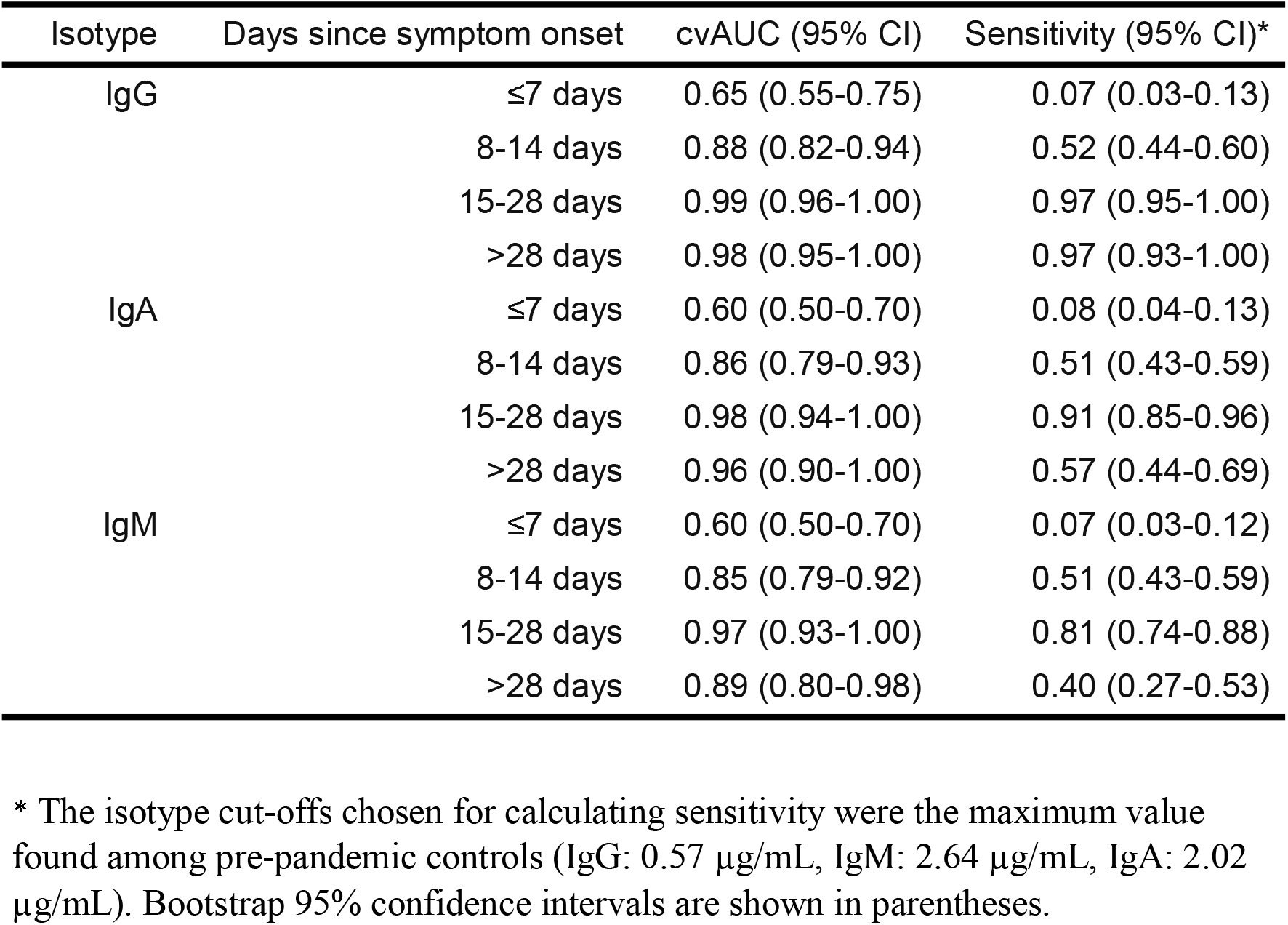
Predictive accuracy of individual isotypes for classifying controls and cases across time.

### Combining multiple isotype measurements to improve predictive accuracy

The discriminatory power of IgG alone was high after 14 days; however, we found the accuracy of ELISA-based identification of recent infections could be improved by adding IgM and/or IgA at the earlier phases of infection (Table S1; Figure S4). The random forest model cvAUC was 0.91 for IgG and IgM and 0.91 for IgG and IgA at 8-14 days post-symptom onset. These models provide an estimate of the contribution of each antibody isotype, as well as the maximum predictive value of combined measures of anti-RBD IgG, IgA and IgM responses. While IgM levels contributed the most to prediction of recent infection in the early phase of illness, IgG responses were the most predictive of infection 8 or more days after the onset of symptoms. Using the pre-determined thresholds for seropositivity for each antibody isotype, out of the 223 patients with samples collected during early infection (< 14 days post symptom onset), we were able to identify an additional 14 cases by adding IgM, 15 by adding IgA, and 23 by adding both IgM and IgA.

### Estimation of time to seroconversion and seroreversion for each isotype

Of 94 cases with samples after 20 days post-symptoms, most had evidence of seroconversion for all isotypes (IgG: 99%, IgM: 90%, IgA: 96%). The estimated median time to seroconversion from symptom onset was comparable across antibody isotype: 10.9 days (95% CI:10.0-11.9) for IgG, 11.8 days (10.7- 13.1) for IgA and 12.1 (10.8-13.7 days) for IgM (Figure 2). On average, most hospitalized individuals seroconverted at least two days sooner to all isotypes compared to non-hospitalized patients, while age and sex appeared to have little effect (Table S2). Of seroconverted cases with samples 40 days post-symptoms or after, most eventually had IgM (21/27) and IgA (18/27) seronegative measurements. Only 1 of 29 (an immunosuppressed individual) cases seroreverted for IgG. We estimated the median time to seroreversion for IgM was 46.9 days (95% CI: 41.1 – 54.8), with the first 5% seroreverting by 24 days. Similarly, we estimate a median seroreversion time for anti-RBD IgA to be 51.0 days (95% CI: 45.7 – 57.1), with the first 5% seroreverting by 29.8 days (Figure 2).

**Figure 2.**
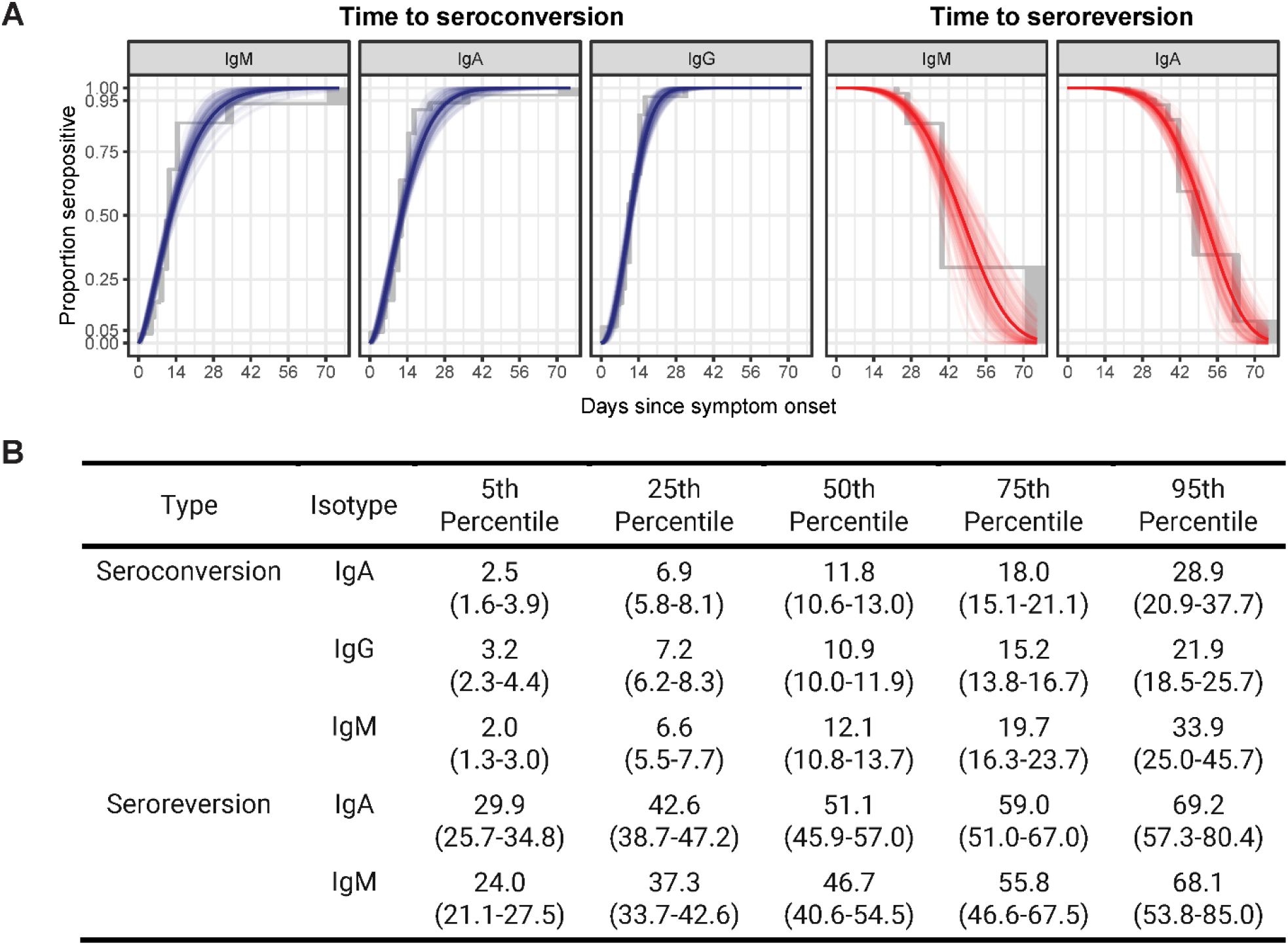
Parametric and non-parametric model estimates of time to seroconversion and seroreversion for each isotype. A) The isotype cut-offs chosen for seroconversion were the maximum concentration (µg/mL) found among pre-pandemic controls (IgG: 0.57, IgM: 2.63, IgA: 2.02). The solid line represents the estimated cumulative distribution function of the time to seroconversion or reversion with 100 bootstrapped fits shown as transparent lines. The parametric accelerated failure time models assume a Weibull time-to-event distribution. Non- parametric estimates shown in grey were calculated using the Turnbull method. Only 1 individual seroreverted for IgG, so no model is included. B) The table indicates the estimated average number of days since onset of symptoms it takes for a percentage of cases to seroconvert or serorevert. Bootstrap 95% confidence intervals are shown in parentheses.

### Association between RBD responses and the development of neutralizing antibodies targeting the S protein

We measured pseudoneutralizing antibodies targeting the SARS-CoV2 S protein in 88 samples from 15 individuals collected between 0 and 75 days post-symptoms (Figure 3). Over the course of infection, all patients tested developed detectable neutralizing antibodies (NAb). NAb titers were correlated with the concentration of anti-RBD IgG (r = 0.87). Of note, similar to anti-RBD IgG responses, NAb titers plateaued and remained detectable at later time points despite the more rapid decline of IgA and IgM responses.

**Figure 3.**
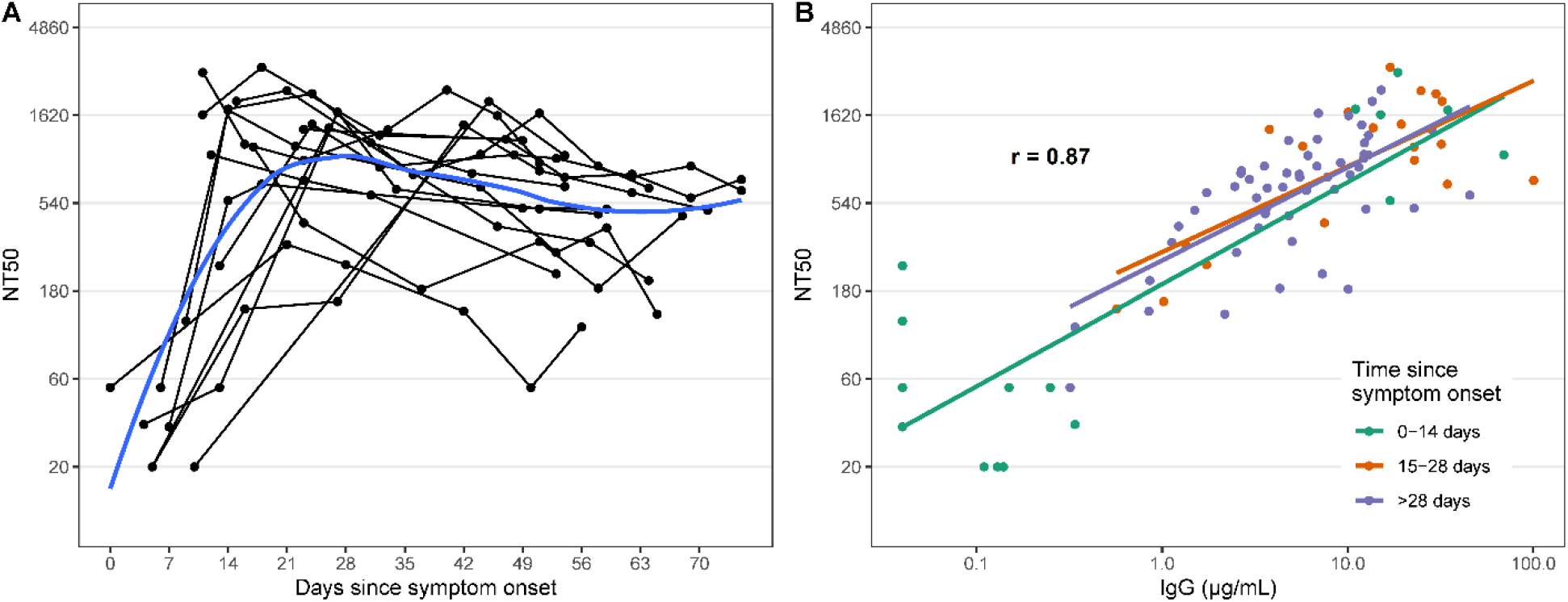
SARS-CoV-2 pseudovirus neutralization antibody titers in symptomatic PCR positive cases and correlation with anti-RBD IgG responses. A) Each point represents a measurement of 50% neutralizing titer (NT50). Lines connect measurements from the same individual and a loess smooth function is shown in blue. B) The overall repeated measures correlation coefficient (r) is shown. Lines represent simple linear models for each time period.

### Evaluation of cross-reactivity with other coronaviruses

We evaluated antibody responses to RBDs derived from spike proteins of endemic human coronaviruses (CoVs) (i.e. HKU1, CoV OC 229E, OC43, and NL63), severe acute respiratory syndrome coronavirus (SARS-CoV-1) and Middle East Respiratory Syndrome coronavirus (MERS-CoV) (Figure S6). We did not observe a cross-reactive response to the endemic human coronaviruses in individuals infected with SARS-CoV-2, and antibody responses to these CoVs were comparable between pre-pandemic controls and individuals with COVID-19 at all phases of infection. We observed significant cross-reactivity to SARS-CoV-1 RBD in individuals with COVID-19, but no cross-reactive responses to the MERS-CoV RBD.

### Comparison of plasma responses to dried blood spots (DBS)

Since this assay could be used in large serosurveys, we also evaluated the assay with dried blood spot eluates in a subset of patients (n= 20 at two timepoints; 40 samples) and pre-pandemic controls (n=20). The anti-RBD IgG DBS measurements had a high degree of linear correlation in both cases and control plasma (r = 0.99, Figure S7). While the classification of all samples was the same between DBS and plasma samples (100% classification concordance), values between the two media diverged more at low titer values.

## DISCUSSION

In this study, we found that the presence of antibodies against the RBD region of the S protein was highly predictive of SARS-CoV-2 infection. Specifically, the presence of IgG antibodies targeting SARS-CoV-2 RBD was a highly specific (100%) and sensitive (97%) marker of infection after 14 days from onset of illness. IgG seropositivity was sustained in patients up to 75 days (last time point measured), and the concentration of these antibodies was highly correlated with pseudovirus NAb titers. In contrast, IgM and IgA responses to RBD were short-lived and with many individuals seroreverting within two months of the onset of illness. Longer term follow-up is needed to determine the duration of IgG responses.

RT-PCR based detection of SARS-CoV-2 is sensitive early in the first week after the onset of symptoms^10^, and our results suggest that the detection of antibodies against the SARS-CoV-2 RBD by ELISA is not likely to contribute significantly to the early diagnosis of COVID- 19. Even models incorporating all isotypes had limited predictive capacity during the first week of illness. However, our results do demonstrate that the detection of IgM and IgA responses improved the sensitivity of serologic testing within two weeks of the onset of symptoms. Beyond two weeks after symptom onset, supplementing viral detection assays with antibody-based testing methods clearly increases sensitivity in diagnosing recent infection^11,12^, particularly as the sensitivity of RT-PCR for SARS-CoV-2 infection wanes^13^. Our results, also, show that the early seroreversion of IgA and IgM responses could be useful in the future in distinguishing previous infection from acute COVID-19. Taken together, these findings suggest specific and clearly defined applications for serologic testing of RBD responses in the clinical setting.

Testing for anti-SARS-CoV-2 RBD antibodies can also be applied in seroepidemiologic studies, even in areas of low prevalence, given their excellent specificity and defined kinetics. Variation in the performance of commercial serologic tests and confusion about the role of antibodies as biomarkers of past infection versus protective immunity has led to widespread misperception that antibody testing may not be accurate or useful^14,15^. In contrast, our study, based on a large sample of cases and controls should provide significant confidence in the potential contribution of serologic measures in public health efforts to identify transmission hot- spots and at-risk populations. In addition, the lack of cross-reactivity of SARS-CoV-2 RBD with common cold coronaviruses should provide additional confidence in the specificity of the assay. Longer term characterization of IgG responses, however, is still needed to assist in design and interpretation of serosurveys.

One notable limitation of our study was that our cohort of individuals with SARS-CoV-2 infection was skewed toward adults with clinically significant disease or with risk factors for disease progression. Individuals with mild or asymptomatic infection have been shown to develop less robust antibody responses^13^, which may lead to false negatives if our proposed assay thresholds are used. Individuals with mild or asymptomatic infection may also serorevert quicker than symptomatic individuals^13^. The gradation of responses by disease severity has been found in other infections, including SARS-CoV-2^13^ and MERS-CoV infection^16^. An association between disease severity and the kinetics of the antibody response is also suggested by our finding that individuals with more severe disease, who required ICU-level care, seroconverted earlier than individuals who did not require ICU-level support.

It is important to note that our assay identifies individuals with recent SARS-CoV-2 infection but does not provide information about whether individuals are protected from subsequent infection. Optimal immunologic correlates of protection for SARS-CoV-2 remain unknown in humans. In many human challenge studies of common cold coronavirus infection, the presence of pre-existing neutralizing antibodies has been associated with protection against the development of symptomatic infection and with decreased viral shedding^17^. In addition, in vaccinated rhesus macaques challenged with SARS-CoV-2 infection, neutralizing antibodies directed at the S protein were also a strong correlate of protective immunity^6^. Thus, neutralization titers, in the absence of other known markers, has become a *de facto* immunologic marker of protection pending further investigation; however, whether there is a threshold titer that is robustly associated with protection remains unclear. Nonetheless, it is notable that anti-RBD IgG antibodies were strongly correlated with the neutralizing antibodies associated with protection in vaccinated macaques^6^. This correlation with neutralizing titers was stronger than observed for other previously tested commercial serologic assays^18^.

Our results, therefore, provide strong support for the application of anti-RBD antibodies as a marker of recent SARS-CoV-2 infection. This approach meets the CDC’s interim guidelines for serologic testing^19^ and has the potential to facilitate accurate diagnosis in clinical settings and the implementation of population-based studies of previous infection globally. While the association between RBD-IgG with neutralizing titers and the persistence of these antibodies at late time points is encouraging, further work is needed to define the optimal antibody mediated correlates of protective immunity.

## Data Availability

We plan to make the data available on Zenodo and at the GitHub repository with the link below after publication.

https://github.com/fjones2222/covid19-serodynamics

## Supplementary Materials

### Method S1. Additional details on laboratory assays

#### Enzyme-linked immunosorbent assay (ELISA)

The RBDs were expressed in Expi293F suspension cells with a C-terminal SBP-His8X tag, and purified using affinity chromatography and then size exclusion chromatography prior to removal of the His tag as described previously^1^. Briefly, 384 well Nunc MaxiSorp plates (Invitrogen, Carlsbad, CA) were coated by adding 50 µL of RBD in carbonate buffer (1 µg/mL) and incubating for 1 hour at room temperature (RT). Plates were then blocked for 30 minutes at RT with 5% non-fat milk in tris-buffered saline (TBS). Diluted samples (1:100 in TBS with 5% milk, 0.5% Tween) were added to the plate (25 µL/well) and incubated for 1 hour at 37°C with shaking. Serial 4-fold dilutions to 1:6400 were also included for individuals with high titers. Goat anti-human IgA, IgG, and IgM-horseradish peroxidase conjugated secondary antibodies diluted (Jackson ImmunoResearch, West Grove, PA) at 1:10000 (IgG, IgM) or 1:5000 (IgA) in 5% milk in TBST were then added to plates (25 µL/well) and incubated at RT with shaking for 30 minutes. Bound secondaries were detected using 1-step Ultra TMB (tetramethylbenzidine; ThermoScientific, Waltham, MA, 25 µL/well). Plates were incubated at RT for 5 minutes in the dark before addition of 2 N sulfuric acid stop solution (25 µL/well). The optical density (OD) was read at 450 nm and 570 nm on a plate reader. OD values were adjusted by subtracting the 570 nm OD from the 450 nm OD. We used a standard curve of the anti-SARS-CoV-2 monoclonal, CR3022^2^,to calculate the concentration of anti-RBD IgG, IgA, and IgM expressed in µg/mL.

#### Dried blood spots (DBS)

Single donor, sero-negative whole blood, collected in sodium heparin tubes (Becton, Dickinson, NJ), was spiked with heat-inactivated plasma from SARS-CoV-2 patients. Forty microliter (40 µL) droplets were spotted in replicate onto Whatman 903 Protein Saver cards (GE Healthcare, Cardiff, UK). Cards were left to dry overnight at ambient room temperature. The following day, two 6-mm^2^ punches were removed from the DBS card using a manual hole-punch and eluted overnight at 4°C with gentle agitation in 133 µL PBS-0.05% Tween 20 pH 7.4 (Sigma-Aldrich, St. Louis, MO).

---

**Figure S1:**
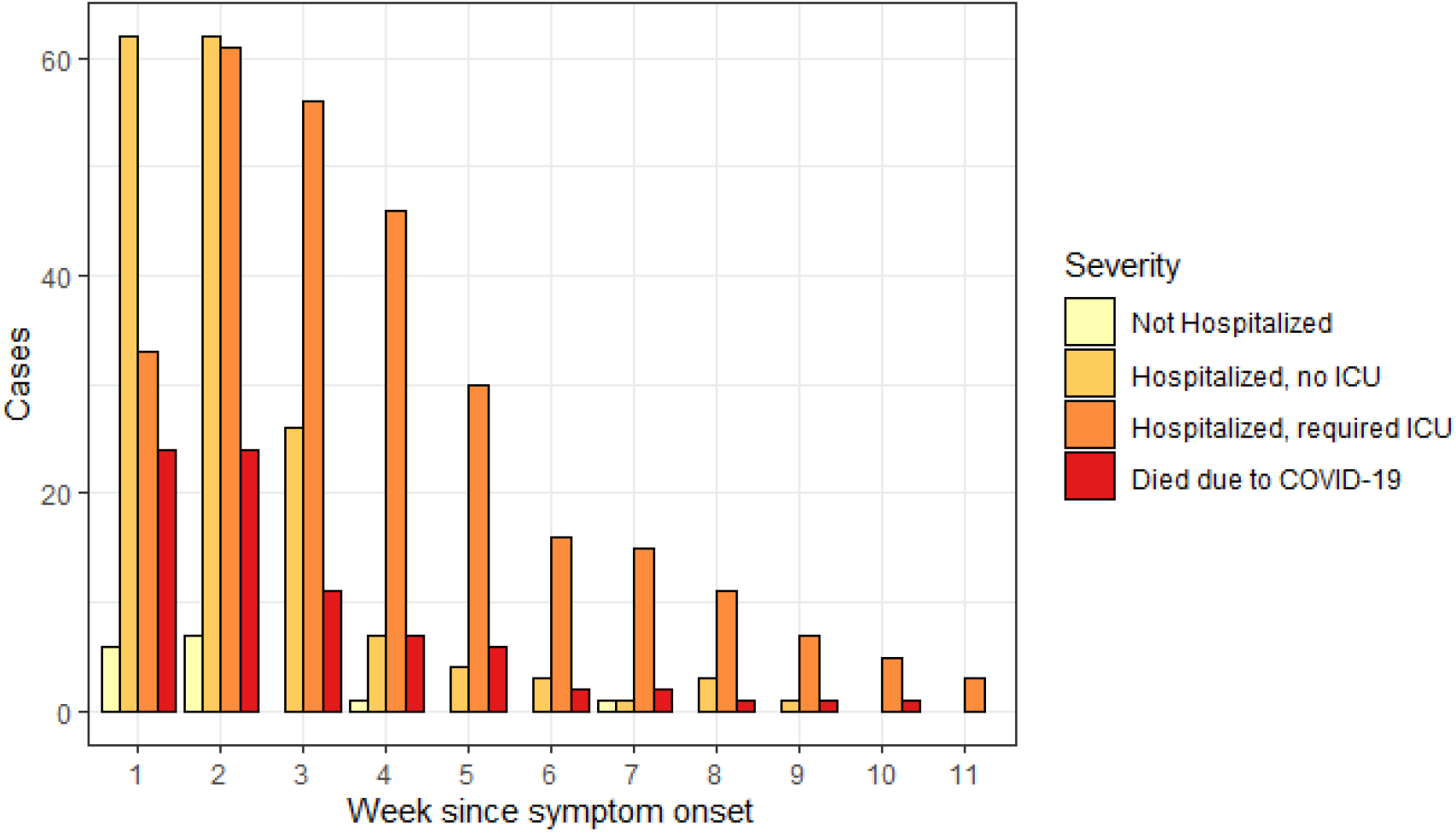
Number of PCR positive cases with a sample taken during each week since symptom onset. The date of symptom onset could not be determined for three individuals and the severity index was missing for one individual

**Figure S2.**
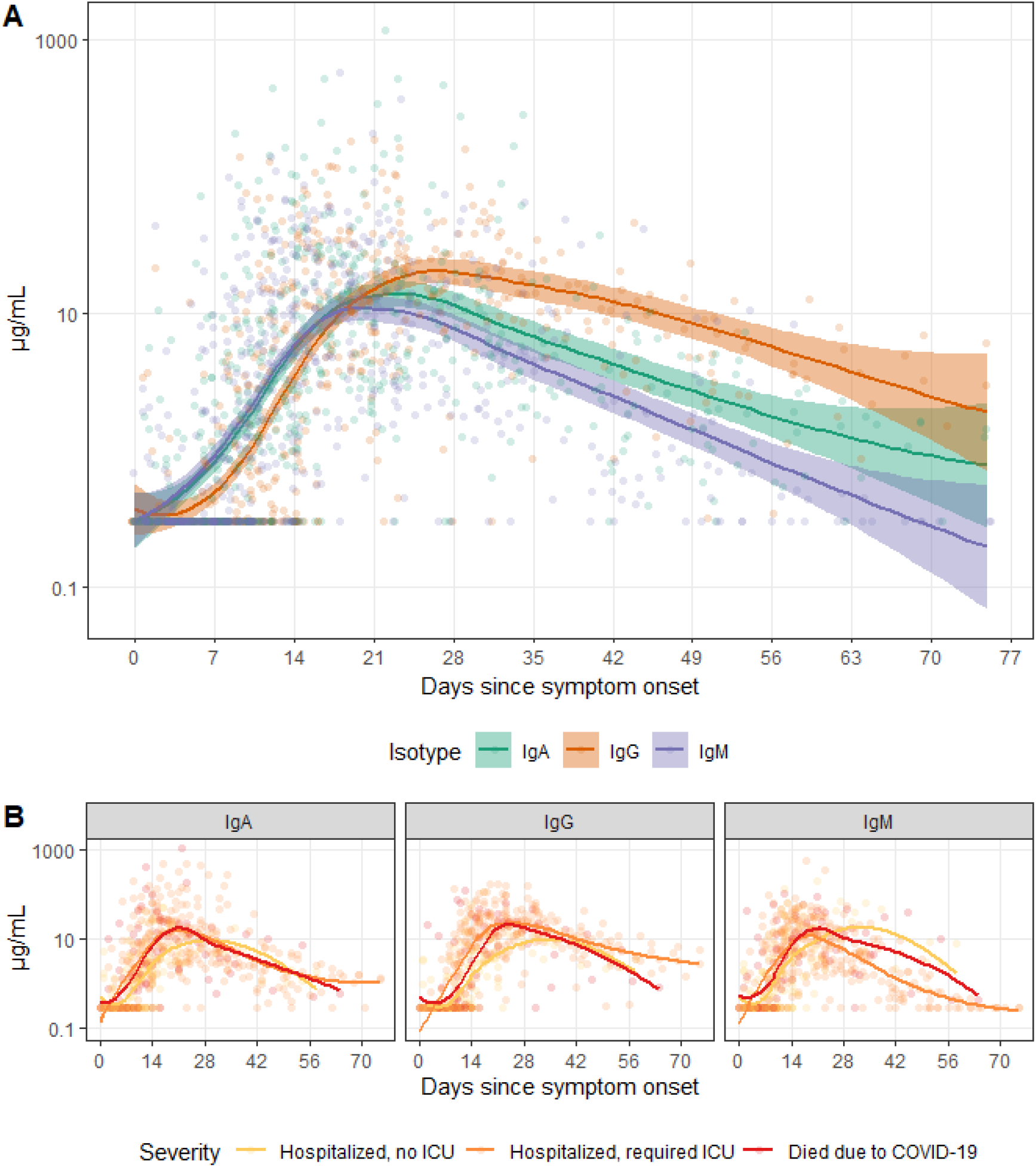
Smooth average measurements of IgG, IgM, and IgA against SARS-CoV-2 spike protein receptor binding domain among PCR positive cases across time. Limit of detection was artificially set at 0.3 µg/mL for IgM and IgG to match that of IgA. Points were jittered horizontally. A) All cases are shown. B) Cases are categorized by clinical severity.

**Figure S3.**
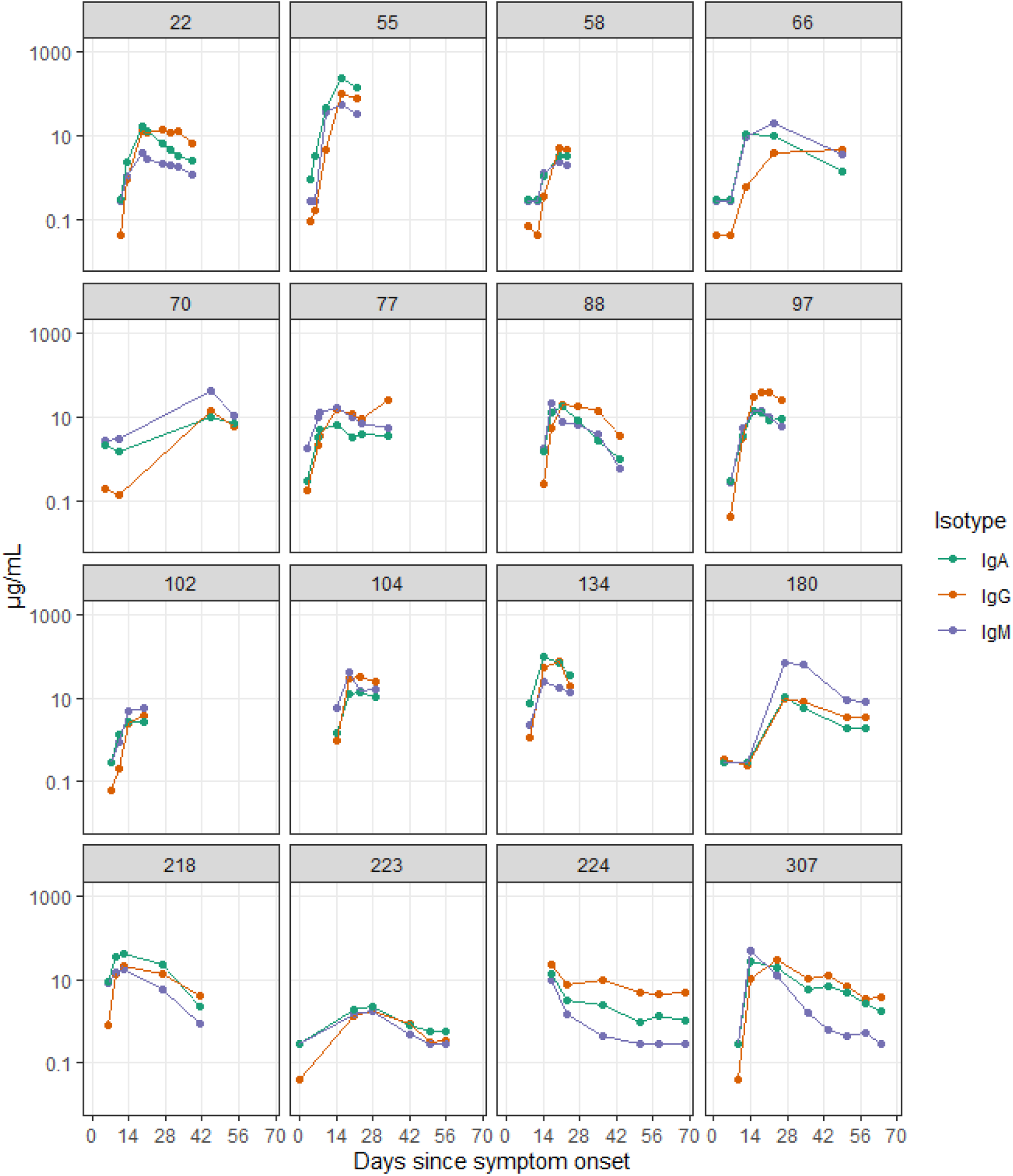
Individual trajectories for 16 randomly selected individuals with 4 or more measurements. Patient ID numbers are shown in greys boxes.

**Table S1.** Predictive accuracy of multiple isotypes for classifying controls and cases over time since symptom onset. Random forest models were used to calculate cvAUC. The isotype cut-offs chosen for calculating sensitivity were the maximum concentration (µg/mL) found among pre-pandemic controls (IgG: 0.57, IgM: 2.63, IgA: 2.02). Samples with measurements above at least one cut-off were classified as cases.

| Isotypes | Days since symptom onset | cvAUC (95% CI) | Sensitivity (95% CI) |
| --- | --- | --- | --- |
| IgA + IgG | $\leq 7$ days | 0.64 (0.54-0.75) | 0.12 (0.09-0.15) |
|  | 8-14 days | 0.91 (0.86-0.96) | 0.55 (0.51-0.59) |
|  | 15-28 days | 0.99 (0.96-1.00) | 0.97 (0.96-0.99) |
| | $> 28$ days | 1.00 (1.00-1.00) | 0.97 (0.96-0.99) |
| IgM + IgG | $\leq 7$ days | 0.64 (0.54-0.74) | 0.10 (0.08-0.13) |
|  | 8-14 days | 0.90 (0.85-0.96) | 0.57 (0.53-0.61) |
|  | 15-28 days | 1.00 (0.99-1.00) | 0.98 (0.96-0.99) |
| | $> 28$ days | 0.98 (0.95-1.00) | 0.97 (0.96-0.99) |
| IgM + IgA | $\leq 7$ days | 0.61 (0.51-0.71) | 0.11 (0.08-0.14) |
|  | 8-14 days | 0.90 (0.84-0.96) | 0.56 (0.51-0.60) |
|  | 15-28 days | 0.99 (0.95-1.00) | 0.95 (0.93-0.97) |
| | $> 28$ days | 0.98 (0.94-1.00) | 0.69 (0.64-0.73) |
| IgM + IgA + IgG | $\leq 7$ days | 0.64 (0.54-0.74) | 0.12 (0.09-0.15) |
|  | 8-14 days | 0.92 (0.86-0.97) | 0.60 (0.56-0.64) |
|  | 15-28 days | 1.00 (0.99-1.00) | 0.98 (0.96-0.99) |
| | $> 28$ days | 1.00 (1.00-1.00) | 0.97 (0.96-0.99) |

**Figure S4.**
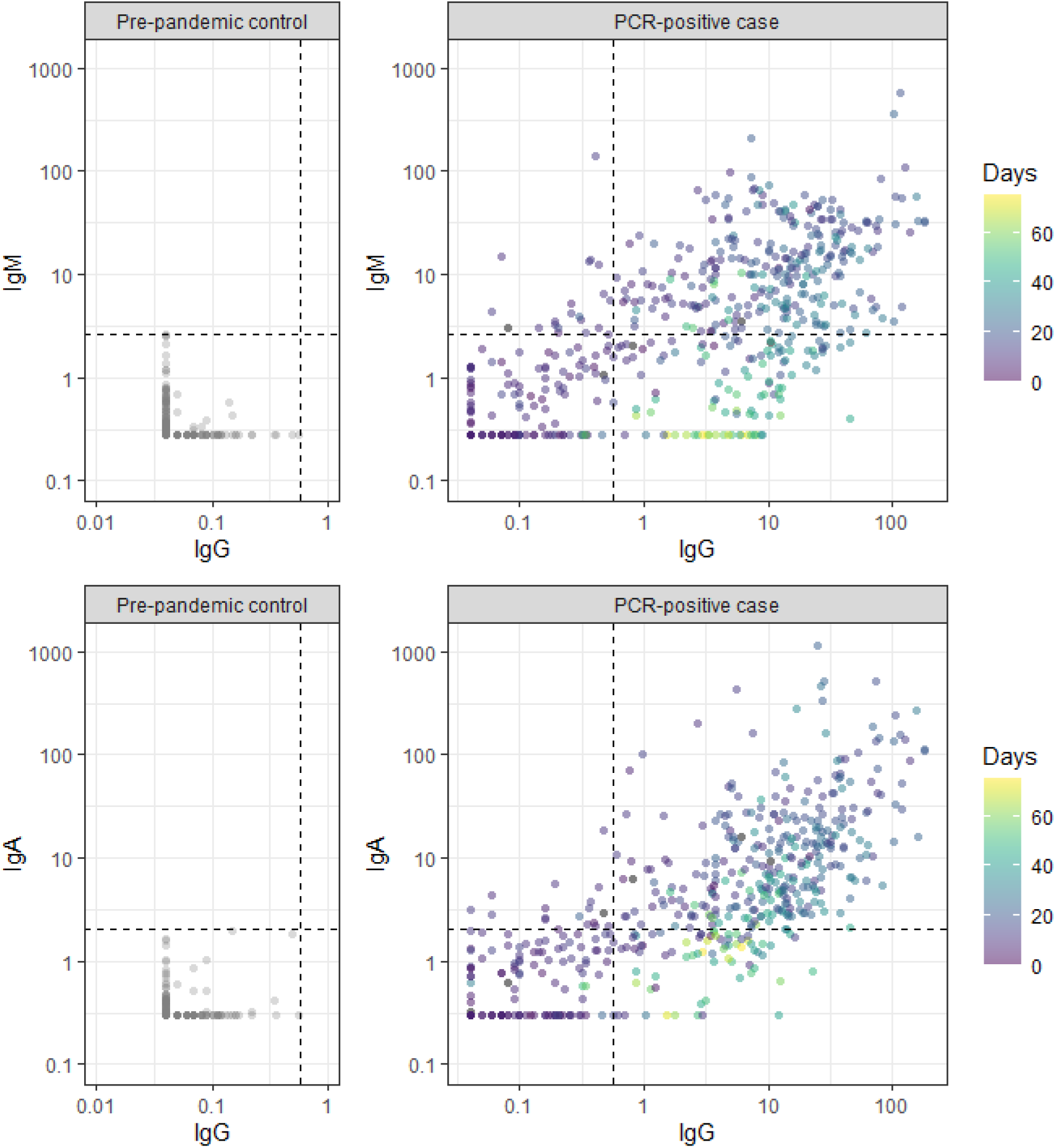
Measurements of IgG, IgM, and IgA against SARS-CoV-2 spike protein receptor binding domain among pre-pandemic controls and symptomatic PCR positive cases. Black dashed line is at 0.57 µg/mL for IgG, 2.63 µg/mL for IgM, and 2.02 µg/mL for IgA.

**Figure S5.**
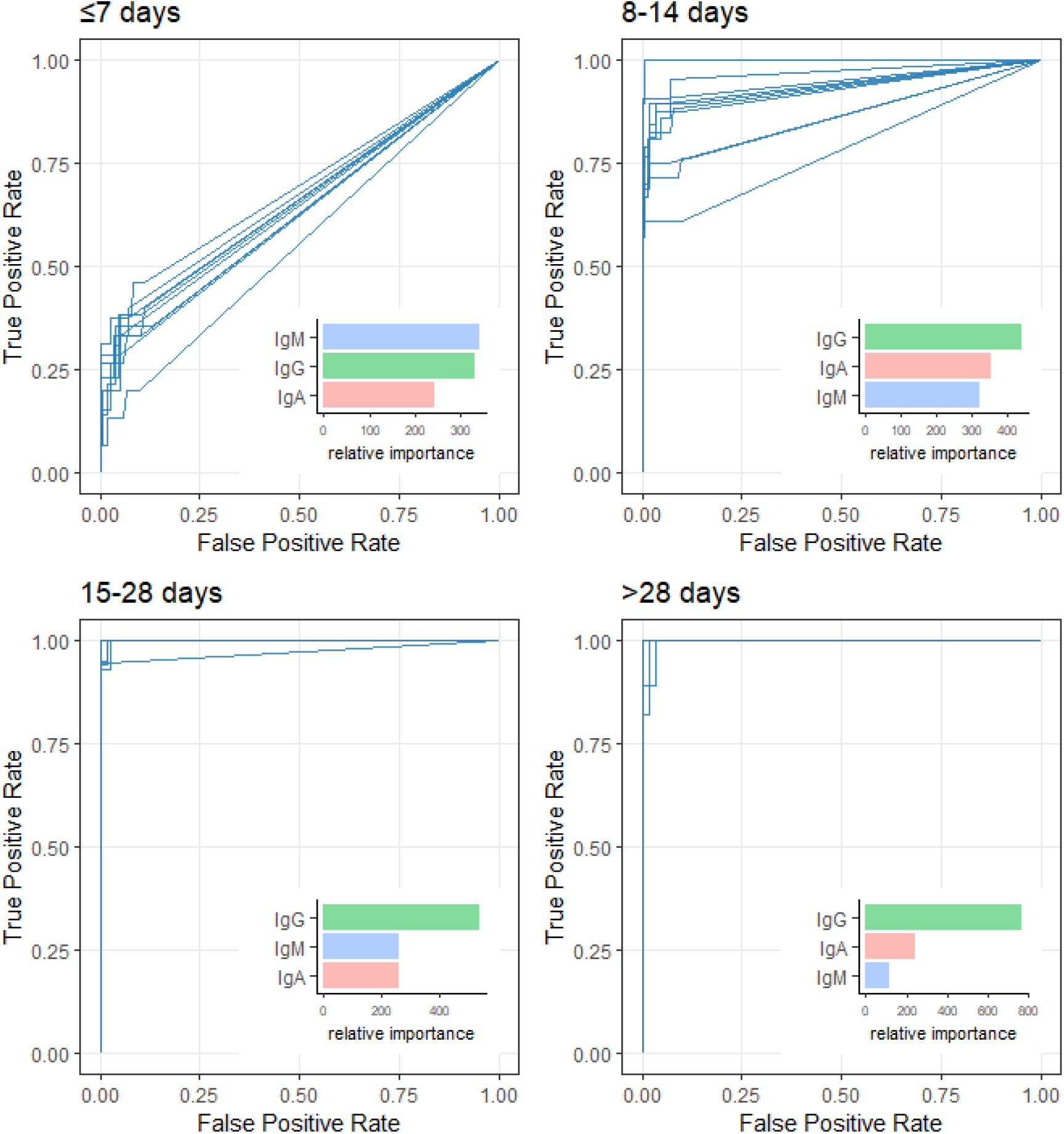
Receiver operating characteristic curve from random forest models and isotype contributions. Each panel shows the ROC curves for cross-validated random forest models fit to serological measurements taken (A) under 7 days (cvAUC: 0.64), (B) 8-14 days (cvAUC: 0.92), (C) 15-28 days (cvAUC: 1.00) and over 28 days (cvAUC: 1.00) after symptom onset of PCR positive cases and pre-pandemic controls. Each blue line is one of ten cross-validated ROC curves for a specific time point. Median relative importance of each serological marker is shown in each bar graph.

**Table S2.** Parametric estimates of median time to seroconversion for each isotype by different patient characteristics. The isotype cut-offs chosen for seroconversion were the maximum concentration (µg/mL) found among pre-pandemic controls (IgG: 0.57, IgM: 2.63, IgA: 2.02). All models assumed that time to event followed a Weibull Distribution. Bootstrap 95% confidence intervals are shown in parentheses. Not Hospitalized and Hospitalized, no ICU were combined due to small sample size.

| Isotype | Characteristic | 50th percentile<br>(95% CI) | Difference<br>(95% CI) |
| --- | --- | --- | --- |
| IgA | Age |  |  |
|  | <65 years | 11.9 (10.7-13.2) |  |
| | $\geq 65$ years | 13.3 (10.2-17.3) | 1.4 (-1.5-5.4) |
|  | Sex |  |  |
|  | Male | 11.8 (10.6-13.1) |  |
|  | Female | 11.5 (9.3-14.1) | -0.3 (-2.6-2.4) |
|  | Severity |  |  |
|  | Not Hospitalized / Hospitalized, no ICU | 12.2 (10.9-13.5) |  |
|  | Hospitalized, required ICU | 9.3 (7.2-11.9) | -2.9 (-5.0--0.4) |
|  | Died due to COVID-19 | 10.1 (7.2-14.1) | -2.1 (-5.0-1.6) |
| IgG | Age |  |  |
|  | <65 years | 10.9 (10.0-11.9) |  |
| | $\geq 65$ years | 12.6 (10.4-15.2) | 1.7 (-0.3-4.0) |
|  | Sex |  |  |
|  | Male | 10.9 (9.9-11.8) |  |
|  | Female | 10.8 (9.2-12.6) | -0.1 (-1.7-1.5) |
|  | Severity |  |  |
|  | Not Hospitalized / Hospitalized, no ICU | 11.3 (10.3-12.3) |  |
|  | Hospitalized, required ICU | 7.4 (6.2-8.8) | -3.8 (-5.2--2.5) |
|  | Died due to COVID-19 | 11.6 (8.4-15.8) | 0.4 (-2.7-4.1) |
| IgM | Age |  |  |
|  | <65 years | 12.1 (10.8-13.7) |  |
| | $\geq 65$ years | 12.4 (9.5-16.0) | 0.3 (-2.6-3.8) |
|  | Sex |  |  |
|  | Male | 12.2 (10.9-13.8) |  |
|  | Female | 16.0 (11.4-22.5) | 3.8 (-0.5-9.9) |
|  | Severity |  |  |
|  | Not Hospitalized / Hospitalized, no ICU | 12.3 (10.8-14.0) |  |
|  | Hospitalized, required ICU | 10.1 (7.4-13.7) | -2.2 (-4.8-1.1) |
|  | Died due to COVID-19 | 9.9 (6.4-14.4) | -2.5 (-5.9-2.0) |

**Figure S6.**
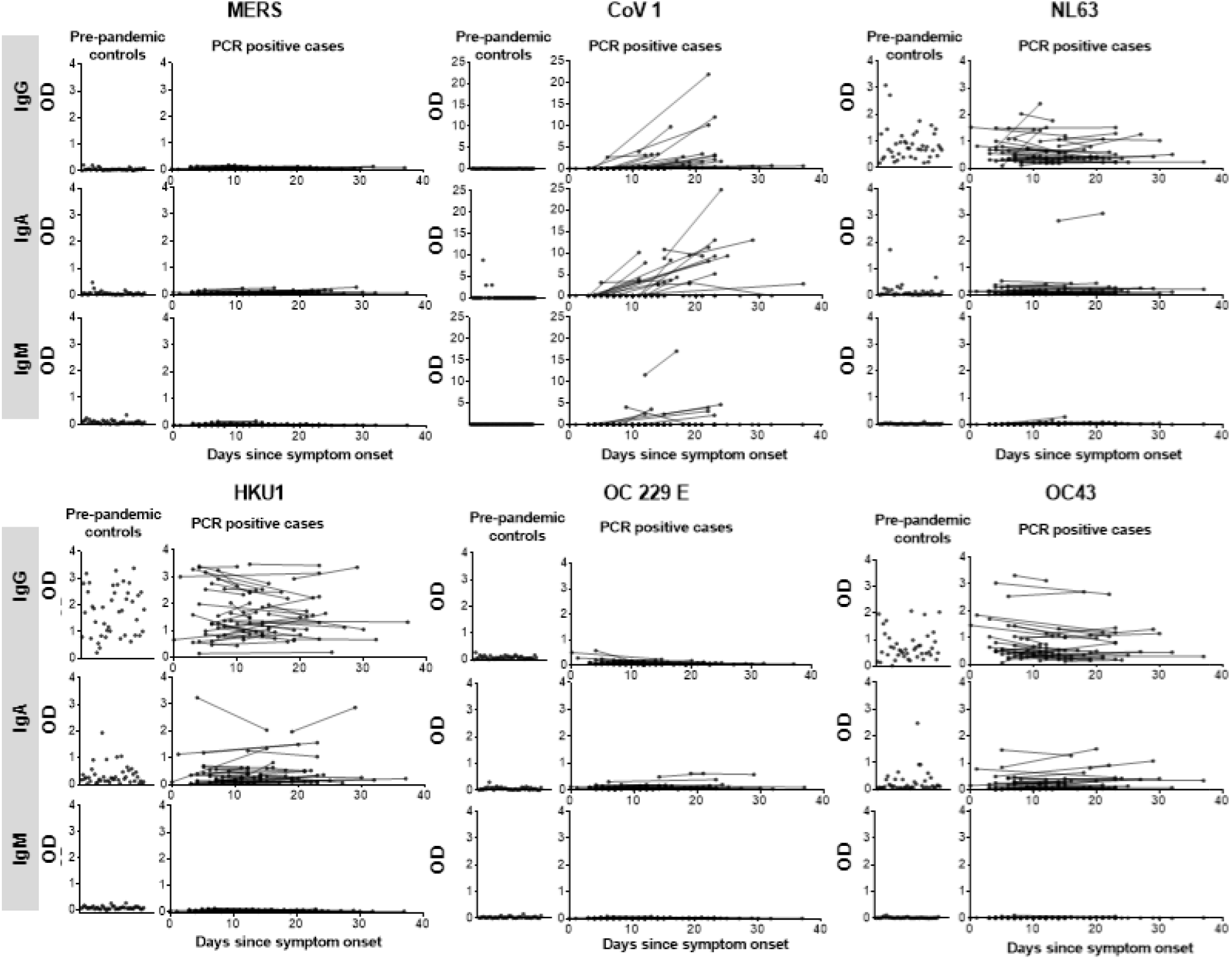
Measurements of IgG, IgA, and IgM against the RBD of other coronaviruses among pre-pandemic controls and PCR positive cases. Each dot represents a unique measurement of a serological marker (Row A: IgG, Row B: IgA, Row C: IgM) in pre-pandemic controls (left panels) and PCR positive cases (right panels) for each coronavirus. Each line connects measurements (dots) for individuals.

**Figure S7.**
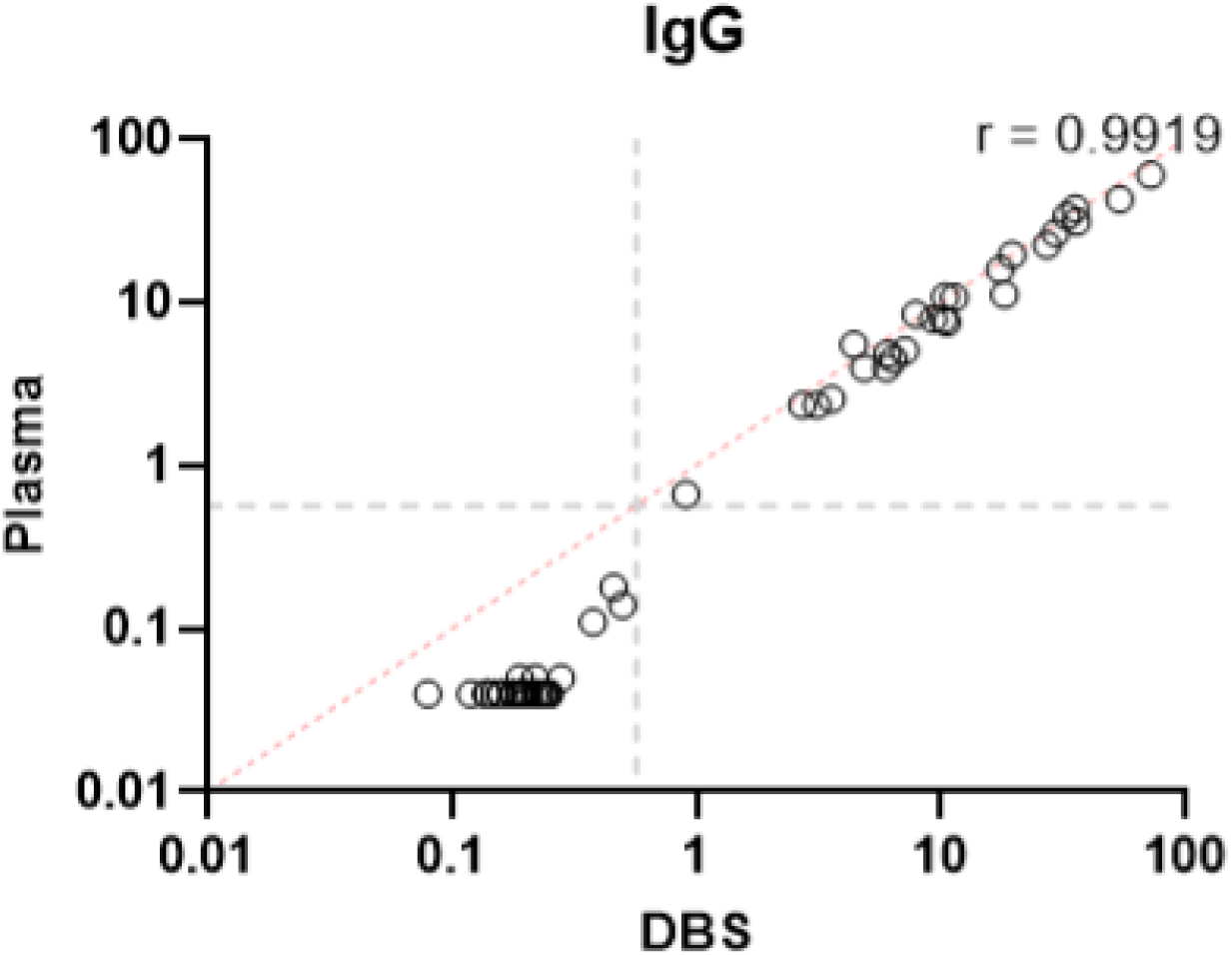
Correlation between plasma and dried blood spot measurements (DBS). Plot of anti-RBD antibody IgG measurement in plasma versus DBS of 20 COVID cases (at 2 timepoints) and 20 pre-pandemic controls. The Pearson correlation coefficient (r) is shown. The dotted gray lines represent the concentration cut-off for seropositivity with plasma.

